# GLP Medications and Severe Post-COVID-19 Outcomes Among Individuals with Type 2 Diabetes Mellitus

**DOI:** 10.64898/2026.07.03.26357246

**Authors:** Zachary Butzin-Dozier, Lin-Chiun Wang, Yunwen Ji, Manav Kumar, A. Jerrod Anzalone, Eric Hurwitz, Rena C. Patel, Ariana Budhihartanto, John B. Buse, Steven Johnson, Carolyn Bramante, Rachel Wong, the National Clinical Cohort Collaborative (N3C) Consortium

## Abstract

**Background:** Glucagon-like peptide-1 receptor agonist-based therapies (GLP) have recently emerged as promising treatments across a wide range of health conditions. These medications may have protective effects against severe long-term consequences of COVID-19 by promoting weight loss, exerting antihyperglycemic and anti-inflammatory effects, and providing cardiovascular and endothelial protection.

**Methods:** We evaluated electronic health record data from a retrospective cohort of individuals in the National Clinical Cohort Collaborative. We included individuals with type 2 diabetes mellitus and comorbid COVID-19 who were prescribed either GLP (treatment) or a sodium-glucose co-transporter 2 inhibitor (SGLT2i) and subsequently developed acute COVID-19 between October 1, 2021, and April 1, 2023. We compared the 12-month cumulative incidence of mortality and Long COVID (Long COVID diagnosis and probable Long COVID via computational phenotype) between groups. We applied targeted maximum likelihood estimation to compare outcome risks by exposure status, controlling for covariates of interest.

**Results:** We analyzed data from 14,215 individuals with COVID-19 and comorbid type 2 diabetes (mean age, 60 years; mean BMI, 37). Compared to SGLT2i, a prescription for GLP medication was associated with a lower risk of mortality (adjusted risk ratio [aRR] 0.71; 95% CI 0.53, 0.95), but not Long COVID diagnosis (aRR 1.01; 95% CI 0.80, 1.27) or probable Long COVID (aRR 0.94; 95% CI 0.88, 1.01).

**Conclusions:** We found that among individuals with type 2 diabetes and comorbid COVID-19, a prescription for GLP vs. SGLT2i medications was associated with a lower risk of mortality, but not Long COVID.

---

Long COVID is a potentially severe syndrome that has impacted one third of the global population.^1^ It is a broad grouping of conditions that individuals experience after acute COVID-19, encompassing symptoms across all biological domains.^2–4^ Type 2 diabetes mellitus (T2DM) is one of the most common metabolic disorders with multiple pathophysiological defects.^5–7^ Individuals with type 2 diabetes are at greater risk of long-term severe sequelae of COVID-19, including mortality or Long COVID.^8–10^ Glucagon-like peptide-1 receptor agonists and combination glucagon-like peptide-1 and glucose-dependent insulinotropic polypeptide dual agonists (hereafter referred to as GLP) may reduce severe long-term sequelae of COVID-19 through weight loss promotion, antihyperglycemic and anti-inflammatory effects, and cardiovascular and endothelial protection.^11,12^ Sodium-glucose co-transporter 2 inhibitors (SGLT2i) are antihyperglycemic medications that reduce blood glucose by increasing urinary glucose excretion and also have demonstrated cardiovascular, renal, and anti-inflammatory benefits.^13^ Both GLP and SGLT2i are widely used therapies for patients with T2DM and share overlapping indications, including promoting weight loss and reducing the risk of adverse cardiovascular and cardiorenal outcomes.^14^ Few studies have evaluated the relationship between GLP medications and Long COVID.

There is evidence that GLP medications are beneficial in preventing adverse outcomes after COVID-19. The 2024 SELECT trial enrolled individuals 45 years of age or older, with a BMI greater than 27 kg/m^2^, cardiovascular disease, and without diabetes, and assigned participants to receive semaglutide (a GLP medication) or a placebo.^15^ The trial found that individuals who developed COVID-19 and were assigned semaglutide had a lower risk of serious COVID-related adverse events (232 vs. 277; *p* = 0.04) and COVID-19-related death (hazard ratio 0.66, 95% CI 0.44-0.96). They also found that among all individuals (including individuals with and without COVID-19), semaglutide was associated with a lower hazard of all-cause mortality (hazard ratio 0.81; 95% CI: 0.71-0.93).^15^

Several observational studies have also provided some insights that prescription of GLP medications versus SGLT2i may be associated with a lower risk of mortality among individuals with T2DM and comorbid COVID-19, but no published studies have directly compared GLP and SGLT2i monotherapy. A cohort study of 78,806 individuals with T2DM and COVID-19 in the National Clinical Cohort Collaborative (N3C) found that GLP therapy was associated with lower 60-day odds of mortality (OR 0.64, 95% CI 0.56-0.72) compared to dipeptidyl peptidase-4 inhibitors (DPP4i), and that the combination GLP and SGLT2i therapy, compared to SGLT2i monotherapy, was not associated with 60-day mortality, although there was a near-significant protective relationship (0.88, 95% CI 0.76 to 1.01).^16^

Although few studies have evaluated the relationship between GLP medications and Long COVID, several studies have evaluated the relationship between GLP medications and COVID-19-related complications. A propensity-matched-cohort study of individuals with T2DM and COVID-19 using data from TriNetX found that prescription for GLP was associated with a lower 28-day cumulative incidence of COVID-19-related respiratory complications (RR 0.62 [95% CI 0.52-0.73]) relative to individuals not prescribed GLP, DPP4i, or pioglitazone.^17^ More broadly, a recent cohort study found that poor glycemic control is associated with an increased risk of Long COVID, supporting the use of antihyperglycemic medications to prevent Long COVID.^18^

While these studies provide modest evidence that GLPs may reduce the risk of mortality and Long COVID, evidence remains limited. In this study, we sought to evaluate the relationship between GLP medications and mortality and Long COVID. We conducted a retrospective cohort study of individuals with T2DM and COVID-19 to compare the 12-month risk of mortality and Long COVID between those prescribed GLP medication and those prescribed SGLT2i.

## METHODS

### Data source

We evaluated data from individuals in N3C with pre-existing T2DM who were diagnosed with COVID-19 between October 1, 2021, and April 1, 2023. This timeframe was chosen to ensure that diagnoses occurred during the Omicron period, after the implementation of the ICD-10 code U09.9 for Long COVID, and to allow for 12 months of follow-up.^19^ We defined T2DM as individuals with either an ICD-10 code for T2DM or a hemoglobin A1C (HbA1C) greater than 6.5%. We excluded individuals who were diagnosed with chronic kidney disease stages 3, 4, or 5, end-stage renal disease, prediabetes, or polycystic ovarian syndrome.^20^ We defined the COVID-19 index date as the earliest date of a diagnosis code (ICD-10-CM U07.1) or a laboratory-confirmed positive result for SARS-CoV-2 infection.^21^

### Exposure

Our exposure of interest was prescription for a GLP medication (semaglutide, dulaglutide, exenatide, liraglutide, lixisenatide, and tirzepatide [glucagon-like peptide-1 and glucose-dependent insulinotropic polypeptide dual agonist]) versus an SGLT2i medication (empagliflozin, dapagliflozin, canagliflozin, ertugliflozin, and bexagliflozin), during acute COVID-19. Given the indications of GLP and SGLT2i as antihyperglycemic medications with anti-inflammatory and cardiometabolic benefits that are commonly prescribed for individuals with T2DM, obesity, or cardiovascular risk factors, SGLT2is are a clinically relevant active comparator frequently used in prior evaluations of GLP therapies.^14,22,23^ Temporally, we considered an individual prescribed a medication (GLP or SGLT2i) if they were prescribed the medication at least 30 days before the documented COVID-19 (index SARS-CoV-2 infection) date, and their prescription end date was not before acute COVID-19. We included prescription data during the N3C observation window beginning in January 2018. We excluded individuals who had been prescribed both GLP and SGLT2i medications within 30 days before COVID-19. We defined treatment as binary, emulating an intention-to-treat randomized trial design; we included only initial prescriptions during our study period and did not consider subsequent treatment changes to avoid potential selection bias.^24^

### Outcomes

Our outcomes of interest were 12-month cumulative mortality and 12-month cumulative incidence of Long COVID, using two definitions: Long COVID diagnosis and probable Long COVID. All outcomes were evaluated within 12 months of acute COVID-19. We defined mortality using linked electronic health record (EHR) and administrative census data. Long COVID diagnosis was defined by the ICD-10 diagnostic code U09.9 (Post-COVID-19 condition, unspecified).^19^ We defined probable Long COVID based on the N3C Long COVID computational phenotype, a computed score of Long COVID probability derived from multiple patient conditions, measurements, and diagnoses available in the EHR.^25–28^ We dichotomized probable Long COVID as present if an individual had a score greater than 0.9 at any time during the 12 months following acute COVID-19, consistent with previous studies.^29,30^

### Covariates

We assessed individuals’ baseline covariate status at the first date of GLP or SGLT2i prescription. For medications and health conditions, we evaluated whether the individuals had any history of use or diagnosis during the N3C observation window, from January 2018 to baseline. For biomarker measurements and body mass index (BMI), we selected the most recent measurement before baseline. For healthcare utilization, we assessed 1) the average healthcare utilization rate (healthcare interactions per month from January 2018 to baseline) and 2) whether the individual had at least 1 healthcare interaction every 6 months in the 12 months after baseline. We adjusted for the following data in our model: average healthcare utilization rate, sex, age, race, data provider, BMI, tobacco use, medical conditions (obesity, chronic lung disease, hypertension, asthma, heart failure, dementia, arthritis, coronary artery disease, cancer, liver disease, chronic kidney disease, peripheral vascular disease, cerebrovascular disease, polycystic ovarian syndrome, and depression), medication use (metformin, systemic corticosteroids, outpatient insulin, angiotensin converting enzyme inhibitors, angiotensin receptor blockers, statins, anticoagulants, aspirin, torsemide, and furosemide), and biomarker measurements (Hba1c, serum creatinine, urine albumin to creatinine ratio, and estimated glomerular filtration rate reported as measurements in the EHR).^31^

### Analysis

We used Super Learner and targeted maximum likelihood estimation to estimate the relationship between GLP prescription vs. SGLT2i prescription, and severe post-COVID-19 outcomes. Our analysis approach incorporates (1) the outcome regression, (2) the treatment mechanism [i.e., inverse probability of treatment], and (3) the censoring mechanism [i.e., inverse probability of censoring] in our causal model.^32–38^ This approach seeks to account for potential biases and confounding present in observational EHR data related to heterogeneous monitoring, difficult-to-characterize treatment mechanisms such as prescribing patterns, and high-dimensional confounding. Super Learner is an ensemble machine learning algorithm, and our candidate learner library included generalized linear models (“SL.glm”), GLM net (“SL.glmnet”), and XGBoost (“SL.xgboost”).^37^ Super Learner is particularly well-suited to this EHR data setting, which includes a wide range of individuals’ covariate information. As traditional parametric models rely on strict assumptions about covariate relationships, we used the Super Learner approach, which combines multiple algorithms to flexibly model these relationships and reduce the risk of misspecification.^36–38^ We used targeted maximum likelihood estimation, a doubly robust method, to estimate risk ratios comparing the 12-month cumulative incidence of our outcomes while adjusting for multiple covariates.^32–36^ Treatment choice between GLP and SGLT2i is strongly influenced by a range of difficult-to-characterize patient and provider characteristics, which can lead to extreme weights, positivity violations, and instability in methods such as inverse probability of treatment weighting. Targeted maximum likelihood estimation is more robust to extreme weights and imperfect characterizations of the treatment mechanism.^32–34,36^ We considered an individual lost to follow-up by censoring (informative right censoring) if/when they had no healthcare interactions during the 12-month follow-up period.^37–39^ For Long COVID-related outcomes, we also considered an individual as informatively censored if/when they died during the 12-month follow-up period. Our analytic approach explicitly models the censoring mechanism (i.e., the inverse probability of censoring) in our causal parameter and evaluates the counterfactual relationship between exposure and outcome under universal monitoring, thereby minimizing bias from heterogeneous monitoring.^37–39^

## RESULTS

We analyzed EHR data from 14,215 individuals in N3C with COVID-19 and comorbid T2DM, of whom 5,130 (60% female) were prescribed GLPs and 9,085 (53% female) were prescribed SGLT2is (Table 1). The mean age of GLP users was 57 years (standard deviation [SD] 11), and that of SGLT2i users was 62 years (SD 12). The average BMI of individuals prescribed GLPs was 40 (SD 9), while the average BMI of individuals prescribed SGLT2is was 35 (SD 8). Individuals prescribed GLPs had, on average, 3.6 healthcare interactions per month (SD 3.9), while those prescribed SGLT2is had, on average, 2.8 healthcare interactions per month (SD 3.3).

**Table 1.** Characteristics of sample individuals with T2DM prescribed GLP vs SGLT2i during acute COVID-19.

| Characteristic | Value | GLP Agonists: *n* (%) | SGLT2i: *n* (%) | Total: *n* (%) |
| --- | --- | --- | --- | --- |
| **Total** |  | 5130 (36%) | 9085 (64%) | 14215 (100%) |
| **Sex** | Female | 3065 (60%) | 4789 (53%) | 7854 (55%) |
| **Age: mean (SD)** |  | 57.05 (11.37) | 62.1 (11.69) | 60.27 (11.83) |
| **Ethnicity** | White Non-Hispanic | 3444 (67%) | 5442 (60%) | 8886 (63%) |
|  | Black or African American Non-Hispanic | 826 (16%) | 1453 (16%) | 2279 (16%) |
|  | Hispanic or Latino Any Race | 434 (8%) | 1066 (12%) | 1500 (11%) |
|  | Unknown | 183 (4%) | 356 (4%) | 539 (4%) |
|  | Asian or Pacific Islander Non-Hispanic | 208 (4%) | 688 (8%) | 896 (6%) |
| **BMI: mean (SD)** |  | 39.77 (9.28) | 35.2 (8.39) | 36.75 (8.97) |
| **Medical Conditions** | Tobacco Smoker | 767 (15%) | 1412 (16%) | 2179 (15%) |
|  | Obese | 4016 (78%) | 5837 (64%) | 9853 (69%) |
|  | Chronic Lung Disease | 1258 (25%) | 2301 (25%) | 3559 (25%) |
|  | Hypertension | 3889 (76%) | 6840 (75%) | 10729 (75%) |
|  | Asthma | 917 (18%) | 1386 (15%) | 2303 (16%) |
|  | Heart Failure | 542 (11%) | 1076 (12%) | 1618 (11%) |
|  | Dementia | 45 (1%) | 191 (2%) | 236 (2%) |
|  | Arthritis | 107 (2%) | 206 (2%) | 313 (2%) |
|  | Coronary Artery Disease | 872 (17%) | 1767 (19%) | 2639 (19%) |
|  | Cancer | 454 (9%) | 1067 (12%) | 1521 (11%) |
|  | Liver Disease | 55 (1%) | 133 (1%) | 188 (1%) |
|  | Chronic Kidney Disease | 417 (8%) | 883 (10%) | 1300 (9%) |
|  | Peripheral Vascular Disease | 531 (10%) | 1181 (13%) | 1712 (12%) |
|  | Cerebrovascular Disease | 346 (7%) | 796 (9%) | 1142 (8%) |
|  | Polycystic Ovarian Syndrome | 138 (3%) | 68 (1%) | 206 (1%) |
|  | Autoimmune Condition | 118 (2%) | 258 (3%) | 376 (3%) |
|  | Immunosuppressed | <20 (<20) | <20 (<20) | <20 (<20) |
|  | Depression | 1582 (31%) | 2216 (24%) | 3798 (27%) |
| **Medications** | Insulin | 2848 (56%) | 4133 (45%) | 6981 (49%) |
|  | ACE Inhibitors | 1296 (25%) | 911 (10%) | 2207 (16%) |
|  | Angiotensin Receptor Blockers | 787 (15%) | 564 (6%) | 1351 (10%) |
|  | Statins | 1708 (33%) | 1306 (14%) | 3014 (21%) |
|  | Anticoagulants | 825 (16%) | 1057 (12%) | 1882 (13%) |
|  | Aspirin | 1151 (22%) | 1166 (13%) | 2317 (16%) |
|  | Systemic Corticosteroids | 2798 (55%) | 4171 (46%) | 6969 (49%) |
|  | Torsemide | 50 (1%) | 116 (1%) | 166 (1%) |
|  | Furosemide | 661 (13%) | 1149 (13%) | 1810 (13%) |
| **Biomarkers** | Pre-Prescription HbA1c: mean (SD) | 8.01 (1.92) | 8.03 (1.79) | 8.02 (1.84) |
|  | Serum Creatinine: mean (SD) | 0.72 (0.4) | 0.85 (0.32) | 0.8 (0.36) |
|  | Albumin/Creatinine Ratio: mean (SD) | 25.15 (56.42) | 13.51 (35.93) | 18.08 (45.45) |
|  | Estimated Glomerular Filtration Rate (eGFR): mean (SD) | 72.92 (29.29) | 70.52 (27.6) | 70.7 (27.74) |
| **Healthcare Utilization** | Interactions per Month: mean (SD) | 3.59 (3.87) | 2.81 (3.25) | 3.09 (3.51) |

We found that individuals with T2DM prescribed GLP medications during acute COVID-19 had a significantly lower 12-month cumulative mortality than individuals prescribed SGLT2i medications (adjusted risk ratio (aRR) 0.71, 95% CI 0.53 to 0.95; unadjusted risk ratio (uRR) 0.44, 95% CI (0.35 to 0.55)) (Figure 1, Table 2). We did not detect a significant relationship between GLP prescription versus SGLT2i prescription and Long COVID via Long COVID diagnosis (aRR 1.01, 95% CI 0.80 to 1.27; uRR 1.01, 95% CI 0.80 to 1.27) or probable Long COVID (aRR 0.94, 95% CI 0.88 to 1.01; uRR 0.94, 95% CI 0.88 to 1.01).

**Figure 1.**
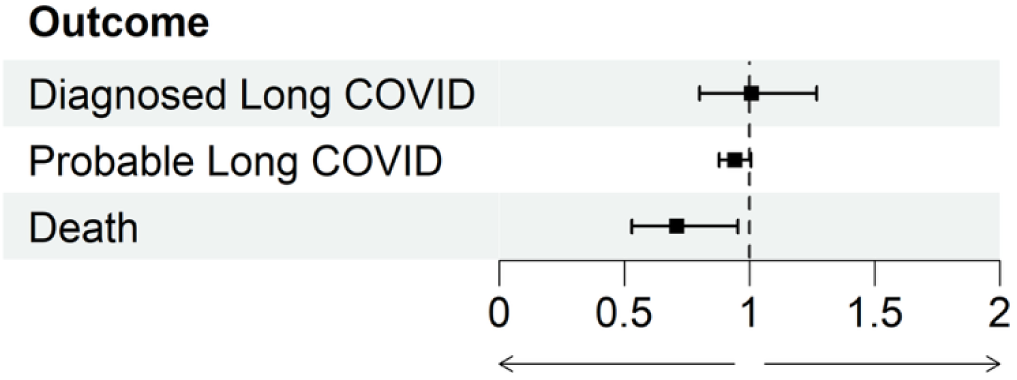
Adjusted risk ratios and 95% confidence intervals describing the relationships between the prescription of GLP (*n* = 5,130) versus SGLT2i (*n* = 9,085) acute COVID-19 and the subsequent 12-month cumulative incidence of mortality and Long COVID outcomes, among individuals with T2DM.

**Table 2.** Unadjusted relationships between the prescription of GLP vs SGLT2i during acute COVID-19 and the subsequent 12-month cumulative incidence of mortality and Long COVID outcomes, among individuals with T2DM.

| Outcome | GLP sample size | SGLT2i sample size | GLP Number of Outcomes | SGLT2i Number of Outcomes | GLP Unadjusted Risk | SGLT2i Unadjusted Risk | Unadjusted RR (95% CI) | Adjusted RR (95% CI) |
| --- | --- | --- | --- | --- | --- | --- | --- | --- |
| Diagnosed Long COVID | 5130 | 9085 | 97 | 142 | 0.02 | 0.02 | 1.21 (0.94 to 1.56) | 1.01 (0.80 to 1.27) |
| Probable Long COVID | 5130 | 9085 | 1095 | 1658 | 0.21 | 0.18 | 1.17 (1.09 to 1.25) | 0.94 (0.88 to 1.01) |
| Death | 5130 | 9085 | 92 | 370 | 0.02 | 0.04 | 0.44 (0.35 to 0.55) | 0.71 (0.53 to 0.95) |

## DISCUSSION

Among individuals with COVID-19 and comorbid T2DM, we found that a prevalent prescription for a GLP medication, compared to SGLT2i, was associated with a lower risk of subsequent mortality but not Long COVID. Given the ubiquity of COVID-19 and the global prevalence of T2DM, this provides important clinical insights regarding the utility of GLP medications to protect a population that is particularly vulnerable to mortality following COVID-19. Our observed effect estimate (aRR = 0.71) is consistent with the findings of the SELECT trial (HR 0.66), supporting the generalizability of these findings in large real-world observational data.^15^ Our findings are also broadly consistent with observational studies showing reduced risk of mortality with GLP compared to non-use or DPP4i, with reported relative point estimates ranging from 0.54 to 0.94.^15,17,20,40^ Cumulatively, these results support that prescription of GLP medications in individuals with T2DM and COVID-19 may be associated with a lower risk of mortality, compared to SGLT2i medications.

We did not detect a significant association between prescription for GLP versus SGLT2i and the subsequent risk of Long COVID. This finding contrasts with a previous study that reported benefits of GLPs for respiratory complications after COVID-19.^17^ The contrast in results is likely due to differences in outcome definition.^41^ Our findings show that while GLP medications may have protective benefits against mortality after COVID-19, there was no evidence to support their clinical use as a preventive pharmacotherapy for Long COVID. Alternatively, SGLT2i may similarly reduce the risk of Long COVID. Given our rapidly developing understanding of the mechanisms and utility of GLP medications, further information on outcomes for which they may not be beneficial helps limit the scope of their applications.

These findings support our understanding of the relationship between GLP medication prescription and subsequent negative health outcomes within a population of patients with COVID-19 and comorbid T2DM. This population represents an intersection of patients with viral infections (COVID-19) and those commonly prescribed GLP medications (T2DM). Our finding of a protective association with mortality supports the wide-ranging benefits of GLP medications, which may be mediated by weight loss, glycemic control, inflammation, and the cardiovascular/endothelial systems, although these mechanisms did not appear to confer protection against Long COVID.^11,12^

### Limitations and strengths

The limited clinical uptake of the Long COVID diagnosis code is a limitation of this study. While many studies with active outcome ascertainment for Long COVID have reported incidence rates exceeding 10%, EHR databases typically report much lower incidence, with our sample showing approximately 2% 12-month cumulative incidence.^3,42^ We sought to address this limitation by including probable Long COVID as an additional outcome with greater sensitivity. Potential exposure misclassification is another limitation. Our study included individual GLP and SGLT2i prescription information, but we were unable to confirm patient receipt or adherence to these prescriptions. Therefore, there is potential for misclassification of true exposure status to GLP and SGLT2i medications, which could be exacerbated by differences in medication costs and delivery methods (i.e., oral versus injectable). The generalizability of N3C is another limitation, as N3C oversamples individuals with high healthcare utilization (i.e., high socioeconomic status, more comorbidities, older).^25,27,28^ By sampling patients with COVID-19, our study implicitly assumes that GLP use is not associated with acute COVID-19 incidence. While we adjusted for many covariates of interest related to individual history and healthcare utilization, it remains possible that residual confounding may distort our estimated relationships.

Major strengths of this study include the data source and the analysis method. The data source, N3C, includes high-dimensional EHR from millions of individuals, enabling the evaluation of rarely diagnosed conditions like Long COVID.^43^ Furthermore, N3C is updated biweekly, which allows for nuanced analyses of recent treatments that might be subject to temporal confounding, such as GLP medications where there have been dramatic changes in utilization patterns during our study period. The analysis method, including Super Learner and targeted maximum likelihood estimation, avoids model misspecification and is robust to near-positivity violations, which are crucial for valid inference in this data setting.^32–34,36^

## CONCLUSIONS

We found that prescription of GLP medications during acute COVID-19 was associated with a lower risk of mortality, compared to SGLT2i medications, among individuals with type 2 diabetes, but we did not detect a similar association with Long COVID.

## Data Availability

All analytic code is available upon request from the N3C Enclave. Access to study data may be requested in the N3C Enclave as legacy data pending N3C approval. Access to the N3C Data Enclave is managed by NCATS. Interested researchers must first complete a data use agreement and next a data use request in order to access the N3C Data Enclave. Once access is granted, the N3C data use committee must review and approve all use of data, and the publication committee must approve all publications involving N3C data.

https://ncats.nih.gov/research/research-activities/n3c/resources/data-access

## DECLARATIONS

### Ethics approval and consent to participate

This study was approved by the UC Berkeley Office for Protection of Human Subjects (2022-01-14980). The N3C data transfer to NCATS is performed under a Johns Hopkins University Reliance Protocol # IRB00249128 or individual site agreements with NIH. N3C received a waiver of consent from the NIH Institutional Review board and allows the secondary analysis of these data without additional consent.

### Consent to publish

The authors consent to the publication of this manuscript in its entirety.

Availability of data and materials: All analytic code is available upon request from the N3C Enclave. Access to study data may be requested in the N3C Enclave as “legacy data” pending N3C approval. Access to the N3C Data Enclave is managed by NCATS (https://ncats.nih.gov/research/research-activities/n3c/resources/data-access). Interested researchers must first complete a data use agreement and next a data use request in order to access the N3C Data Enclave. Once access is granted, the N3C data use committee must review and approve all use of data, and the publication committee must approve all publications involving N3C data.

## N3C Attribution

The analyses described in this manuscript were conducted with data or tools accessed through the NCATS N3C Data Enclave (RP-T9ZZW78) https://covid.cd2h.org and N3C Attribution & Publication Policy v 1.2-2020-08-25b supported by NCATS U24 TR002306, Axle Informatics Subcontract: NCATS-P00438-B, the Bill & Melinda Gates Foundation: OPP1165144, and the National Institutes of General Medical Sciences: U54GM115458 and 5U54GM104942-04. This research was possible because of the individuals whose information is included within the data and the organizations (https://ncats.nih.gov/n3c/resources/data-contribution/data-transfer-agreement-signatories) and scientists who have contributed to the ongoing development of this community resource [https://doi.org/10.1093/jamia/ocaa196].

## Disclaimer

The N3C Publication committee confirmed that this manuscript (MSID PENDING) complies with N3C data use and attribution policies; however, the authors are solely responsible for its content, which does not necessarily represent the official views of the National Institutes of Health or the N3C program.

## IRB

The N3C data transfer to NCATS is performed under a Johns Hopkins University Reliance Protocol # IRB00249128 or individual site agreements with NIH. The N3C Data Enclave is managed under the authority of the NIH; information can be found at https://ncats.nih.gov/n3c/resources.

This research project was approved by the University of California, Berkeley Committee for the Protection of Human Subjects (CPHS protocol number 2022-01-14980). This approval is issued under University of California, Berkeley Federalwide Assurance #00006252.

## Individual Acknowledgements For Core Contributors

We gratefully acknowledge the following core contributors to N3C:

Adam B. Wilcox, Adam M. Lee, Alexis Graves, Alfred (Jerrod) Anzalone, Amin Manna, Amit Saha, Amy Olex, Andrea Zhou, Andrew E. Williams, Andrew Southerland, Andrew T. Girvin, Anita Walden, Anjali A. Sharathkumar, Benjamin Amor, Benjamin Bates, Brian Hendricks, Brijesh Patel, Caleb Alexander, Carolyn Bramante, Cavin Ward-Caviness, Charisse Madlock-Brown, Christine Suver, Christopher Chute, Christopher Dillon, Chunlei Wu, Clare Schmitt, Cliff Takemoto, Dan Housman, Davera Gabriel, David A. Eichmann, Diego Mazzotti, Don Brown, Eilis Boudreau, Elaine Hill, Elizabeth Zampino, Emily Carlson Marti, Emily R. Pfaff, Evan French, Farrukh M Koraishy, Federico Mariona, Fred Prior, George Sokos, Greg Martin, Harold Lehmann, Heidi Spratt, Hemalkumar Mehta, Hongfang Liu, Hythem Sidky, J.W. Awori Hayanga, Jami Pincavitch, Jaylyn Clark, Jeremy Richard Harper, Jessica Islam, Jin Ge, Joel Gagnier, Joel H. Saltz, Joel Saltz, Johanna Loomba, John Buse, Jomol Mathew, Joni L. Rutter, Julie A. McMurry, Justin Guinney, Justin Starren, Karen Crowley, Katie Rebecca Bradwell, Kellie M. Walters, Ken Wilkins, Kenneth R. Gersing, Kenrick Dwain Cato, Kimberly Murray, Kristin Kostka, Lavance Northington, Lee Allan Pyles, Leonie Misquitta, Lesley Cottrell, Lili Portilla, Mariam Deacy, Mark M. Bissell, Marshall Clark, Mary Emmett, Mary Morrison Saltz, Matvey B. Palchuk, Melissa A. Haendel, Meredith Adams, Meredith Temple-O’Connor, Michael G. Kurilla, Michele Morris, Nabeel Qureshi, Nasia Safdar, Nicole Garbarini, Noha Sharafeldin, Ofer Sadan, Patricia A. Francis, Penny Wung Burgoon, Peter Robinson, Philip R.O. Payne, Rafael Fuentes, Randeep Jawa, Rebecca Erwin-Cohen, Rena Patel, Richard A. Moffitt, Richard L. Zhu, Rishi Kamaleswaran, Robert Hurley, Robert T. Miller, Saiju Pyarajan, Sam G. Michael, Samuel Bozzette, Sandeep Mallipattu, Satyanarayana Vedula, Scott Chapman, Shawn T. O’Neil, Soko Setoguchi, Stephanie S. Hong, Steve Johnson, Tellen D. Bennett, Tiffany Callahan, Umit Topaloglu, Usman Sheikh, Valery Gordon, Vignesh Subbian, Warren A. Kibbe, Wenndy Hernandez, Will Beasley, Will Cooper, William Hillegass, Xiaohan Tanner Zhang. Details of contributions available at covid.cd2h.org/core-contributors

## Data Partners with Released Data

The following institutions whose data is released or pending:

Available: Advocate Health Care Network — UL1TR002389: The Institute for Translational Medicine (ITM) • Boston University Medical Campus — UL1TR001430: Boston University Clinical and Translational Science Institute • Brown University — U54GM115677: Advance Clinical Translational Research (Advance-CTR) • Carilion Clinic — UL1TR003015: iTHRIV Integrated Translational health Research Institute of Virginia • Charleston Area Medical Center — U54GM104942: West Virginia Clinical and Translational Science Institute (WVCTSI) • Children’s Hospital Colorado — UL1TR002535: Colorado Clinical and Translational Sciences Institute • Columbia University Irving Medical Center — UL1TR001873: Irving Institute for Clinical and Translational Research • Duke University — UL1TR002553: Duke Clinical and Translational Science Institute • George Washington Children’s Research Institute — UL1TR001876: Clinical and Translational Science Institute at Children’s National (CTSA-CN) • George Washington University — UL1TR001876: Clinical and Translational Science Institute at Children’s National (CTSA-CN) • Indiana University School of Medicine — UL1TR002529: Indiana Clinical and Translational Science Institute • Johns Hopkins University — UL1TR003098: Johns Hopkins Institute for Clinical and Translational Research • Loyola Medicine — Loyola University Medical Center • Loyola University Medical Center — UL1TR002389: The Institute for Translational Medicine (ITM) • Maine Medical Center — U54GM115516: Northern New England Clinical & Translational Research (NNE-CTR) Network • Massachusetts General Brigham — UL1TR002541: Harvard Catalyst • Mayo Clinic Rochester — UL1TR002377: Mayo Clinic Center for Clinical and Translational Science (CCaTS) • Medical University of South Carolina — UL1TR001450: South Carolina Clinical & Translational Research Institute (SCTR) • Montefiore Medical Center — UL1TR002556: Institute for Clinical and Translational Research at Einstein and Montefiore • Nemours — U54GM104941: Delaware CTR ACCEL Program • NorthShore University HealthSystem — UL1TR002389: The Institute for Translational Medicine (ITM) • Northwestern University at Chicago — UL1TR001422: Northwestern University Clinical and Translational Science Institute (NUCATS) • OCHIN — INV-018455: Bill and Melinda Gates Foundation grant to Sage Bionetworks • Oregon Health & Science University — UL1TR002369: Oregon Clinical and Translational Research Institute • Penn State Health Milton S. Hershey Medical Center — UL1TR002014: Penn State Clinical and Translational Science Institute • Rush University Medical Center — UL1TR002389: The Institute for Translational Medicine (ITM) • Rutgers, The State University of New Jersey — UL1TR003017: New Jersey Alliance for Clinical and Translational Science • Stony Brook University — U24TR002306 • The Ohio State University — UL1TR002733: Center for Clinical and Translational Science • The State University of New York at Buffalo — UL1TR001412: Clinical and Translational Science Institute • The University of Chicago — UL1TR002389: The Institute for Translational Medicine (ITM) • The University of Iowa — UL1TR002537: Institute for Clinical and Translational Science • The University of Miami Leonard M. Miller School of Medicine — UL1TR002736: University of Miami Clinical and Translational Science Institute • The University of Michigan at Ann Arbor — UL1TR002240: Michigan Institute for Clinical and Health Research • The University of Texas Health Science Center at Houston — UL1TR003167: Center for Clinical and Translational Sciences (CCTS) • The University of Texas Medical Branch at Galveston — UL1TR001439: The Institute for Translational Sciences • The University of Utah — UL1TR002538: Uhealth Center for Clinical and Translational Science • Tufts Medical Center — UL1TR002544: Tufts Clinical and Translational Science Institute • Tulane University — UL1TR003096: Center for Clinical and Translational Science • University Medical Center New Orleans — U54GM104940: Louisiana Clinical and Translational Science (LA CaTS) Center • University of Alabama at Birmingham — UL1TR003096: Center for Clinical and Translational Science • University of Arkansas for Medical Sciences — UL1TR003107: UAMS Translational Research Institute • University of Cincinnati — UL1TR001425: Center for Clinical and Translational Science and Training • University of Colorado Denver, Anschutz Medical Campus — UL1TR002535: Colorado Clinical and Translational Sciences Institute • University of Illinois at Chicago — UL1TR002003: UIC Center for Clinical and Translational Science • University of Kansas Medical Center — UL1TR002366: Frontiers: University of Kansas Clinical and Translational Science Institute • University of Kentucky — UL1TR001998: UK Center for Clinical and Translational Science • University of Massachusetts Medical School Worcester — UL1TR001453: The UMass Center for Clinical and Translational Science (UMCCTS) • University of Minnesota — UL1TR002494: Clinical and Translational Science Institute • University of Mississippi Medical Center — U54GM115428: Mississippi Center for Clinical and Translational Research (CCTR) • University of Nebraska Medical Center — U54GM115458: Great Plains IDeA-Clinical & Translational Research • University of North Carolina at Chapel Hill — UL1TR002489: North Carolina Translational and Clinical Science Institute • University of Oklahoma Health Sciences Center — U54GM104938: Oklahoma Clinical and Translational Science Institute (OCTSI) • University of Rochester — UL1TR002001: UR Clinical & Translational Science Institute • University of Southern California — UL1TR001855: The Southern California Clinical and Translational Science Institute (SC CTSI) • University of Vermont — U54GM115516: Northern New England Clinical & Translational Research (NNE-CTR) Network • University of Virginia — UL1TR003015: iTHRIV Integrated Translational health Research Institute of Virginia • University of Washington — UL1TR002319: Institute of Translational Health Sciences • University of Wisconsin-Madison — UL1TR002373: UW Institute for Clinical and Translational Research • Vanderbilt University Medical Center — UL1TR002243: Vanderbilt Institute for Clinical and Translational Research • Virginia Commonwealth University — UL1TR002649: C. Kenneth and Dianne Wright Center for Clinical and Translational Research • Wake Forest University Health Sciences — UL1TR001420: Wake Forest Clinical and Translational Science Institute • Washington University in St. Louis — UL1TR002345: Institute of Clinical and Translational Sciences • Weill Medical College of Cornell University — UL1TR002384: Weill Cornell Medicine Clinical and Translational Science Center • West Virginia University — U54GM104942: West Virginia Clinical and Translational Science Institute (WVCTSI) Submitted: Icahn School of Medicine at Mount Sinai — UL1TR001433: ConduITS Institute for Translational Sciences • The University of Texas Health Science Center at Tyler — UL1TR003167: Center for Clinical and Translational Sciences (CCTS) • University of California, Davis — UL1TR001860: UCDavis Health Clinical and Translational Science Center • University of California, Irvine — UL1TR001414: The UC Irvine Institute for Clinical and Translational Science (ICTS) • University of California, Los Angeles — UL1TR001881: UCLA Clinical Translational Science Institute • University of California, San Diego — UL1TR001442: Altman Clinical and Translational Research Institute • University of California, San Francisco — UL1TR001872: UCSF Clinical and Translational Science Institute Pending: Arkansas Children’s Hospital — UL1TR003107: UAMS Translational Research Institute • Baylor College of Medicine — None (Voluntary) • Children’s Hospital of Philadelphia — UL1TR001878: Institute for Translational Medicine and Therapeutics • Cincinnati Children’s Hospital Medical Center — UL1TR001425: Center for Clinical and Translational Science and Training • Emory University — UL1TR002378: Georgia Clinical and Translational Science Alliance • HonorHealth — None (Voluntary) • Loyola University Chicago — UL1TR002389: The Institute for Translational Medicine (ITM) • Medical College of Wisconsin — UL1TR001436: Clinical and Translational Science Institute of Southeast Wisconsin • MedStar Health Research Institute — UL1TR001409: The Georgetown-Howard Universities Center for Clinical and Translational Science (GHUCCTS) • MetroHealth — None (Voluntary) • Montana State University — U54GM115371: American Indian/Alaska Native CTR • NYU Langone Medical Center — UL1TR001445: Langone Health’s Clinical and Translational Science Institute • Ochsner Medical Center — U54GM104940: Louisiana Clinical and Translational Science (LA CaTS) Center • Regenstrief Institute — UL1TR002529: Indiana Clinical and Translational Science Institute • Sanford Research — None (Voluntary) • Stanford University — UL1TR003142: Spectrum: The Stanford Center for Clinical and Translational Research and Education • The Rockefeller University — UL1TR001866: Center for Clinical and Translational Science • The Scripps Research Institute — UL1TR002550: Scripps Research Translational Institute • University of Florida — UL1TR001427: UF Clinical and Translational Science Institute • University of New Mexico Health Sciences Center — UL1TR001449: University of New Mexico Clinical and Translational Science Center • University of Texas Health Science Center at San Antonio — UL1TR002645: Institute for Integration of Medicine and Science • Yale New Haven Hospital — UL1TR001863: Yale Center for Clinical Investigation

## CONTRIBUTORS

All authors read and approved the final version of the manuscript.

Authorship was determined using ICMJE recommendations.

ZB: Generated research question, drafted manuscript, managed project timeline, and coordinated analysis.

YJ, LW, JA, RCP, AB, EH, MK, JB, SJ, CB, and RW: Provided oversight on study design and analysis plan, supported analysis, reviewed manuscript, and provided feedback.

ZB, YJ, and LW have accessed and verified the underlying data.

*Inclusion and ethics statement:* All co-authors and collaborators included in this manuscript have fulfilled the criteria for authorship.

